# SARS-CoV-2 testing and COVID-19 related primary care use among people with citizenship, permanent residency, and temporary immigration status in British Columbia: Cross-sectional analysis of population-based administrative data

**DOI:** 10.1101/2021.11.05.21265978

**Authors:** M Wiedmeyer, S Goldenberg, S Peterson, S Wanigaratne, S Machado, E Tayyar, M Braschel, R Carrillo, C Sierra-Heredia, G Tuyisenge, MR Lavergne

## Abstract

**Background:** Having temporary immigration status affords limited rights, workplace protections, and access to services. There is not yet research data on impacts of the COVID-19 pandemic for people with temporary immigration status in Canada.

**Methods:** We use linked administrative data to describe SARS-CoV-2 testing, positive tests, and COVID-19 primary care service use in British Columbia from January 1, 2020, to July 31, 2021, stratified by immigration status (Citizen, Permanent Resident, Temporary Resident). We plot the rate of people tested and the rate of people confirmed positive for COVID-19 by week from April 19, 2020, to July 31, 2021, across immigration groups.

**Results:** 4.9% of people with temporary immigration status had a positive test for SARS-CoV-2 over this period, compared to 4.0% among people with permanent residency and 2.1% among people who hold Canadian citizenship. This pattern is persistent by sex/gender, age group, neighborhood income quintile, health authority, and in both metropolitan and small urban settings. At the same time we observe lower access to testing and COVID-19 related primary care among people with temporary status.

**Interpretation:** People with temporary immigration status in BC experience higher SARS-CoV-2 test positivity; alarmingly, this was coupled with lower access to testing and primary care. Interwoven immigration, health and occupational policies place people with temporary status in circumstances of precarity and higher health risk. Extending permanent residency status to all immigrants residing in Canada and decoupling access to health care from immigration status could reduce precarity due to temporary immigration status.

## Introduction

Globally, uneven responses to the COVID-19 pandemic have caused disproportionate harm to immigrants, racialized people, and other socially and economically marginalized groups (1–4). COVID-19 outbreaks in places employing immigrants have been widely reported (5–8) and suggest that many people with temporary immigration status have been placed in high risk circumstances (9), though research data is limited. Other people with temporary immigration status, such as international students, have reported serious impacts (10) but are rarely included in government support measures or research (15)(11). Refugee claimants experienced a high burden of COVID-19 (12), despite opportunities to claim asylum being curtailed due to pandemic-related travel restrictions (13–15). An Ontario report in fall 2020 found that while immigrants, refugees and other newcomers made up just 25% of the Ontario population, they experienced 43.5% of all COVID-19 cases (2). These initial findings indicate a relationship between immigration status and circumstances of vulnerability to COVID-19.

Canada regulates immigration through a variety of pathways which can be broadly grouped into temporary or permanent resident status, where permanent residents may become citizens after meeting requirements [Box 1]. The number of temporary permits issued for work or study in Canada has increased dramatically over the past decade (16). Having temporary immigration status affords limited rights, workplace protections, and access to services which together create precarity – a multidimensional insecurity of work, residence, entitlements and health (17–19). This precarity produces harmful health outcomes in people with temporary immigration status in Canada (20–22). We do not yet have research data on the health impact of the COVID-19 pandemic for people with temporary immigration status in Canada, nor data by immigration status outside of Ontario. We use health system data to describe SARS-CoV-2 testing and COVID-19 related primary care by immigration status for people eligible for the provincial insurance plan in the province of British Columbia (BC).

## Methods

### Study setting

The borders of the province of British Columbia (BC) were defined on the lands of more than 200 Indigenous nations through resisted historical and ongoing colonial processes including forced displacement (23). Immigration, Refugees and Citizenship Canada (IRCC) controls movement across federal borders by issuing travel documents and screening potential permanent and temporary residents (24).

The Medical Services Plan (MSP) is BC’s provincial health insurance program that covers health care benefits for eligible BC residents. People who hold Canadian citizenship and people with permanent residency are eligible for MSP provided they meet some conditions (25). Some people who hold temporary study or work permits that are valid for six or more months are also eligible, including refugee claimants and convention refugees who also hold work permits (25). In April 2020, MSP extended temporary coverage for some people with expired work or study permits and established a mechanism to reimburse providers for COVID-19 related care for people without MSP, but not other health services (26). Usually, people who are not eligible, including those whose temporary permits expire, have no access to public health insurance in BC. New eligible residents in BC are required to undergo a 3 month wait period prior to health insurance activation. The province temporarily eliminated this policy in March 2020, only to reinstate it in August 2020.

### Data and study population

We accessed linked, population-based administrative data through Population Data BC, including the MSP registry file (27), SARS-CoV2 testing (28), physician payments (29). Testing, and service use data covers the period from January 1, 2020-July 31, 2021. Analyses included all people registered for MSP at any time January 1, 2020-March 31, 2021. We excluded people identified in the MSP registry as visitors, diplomats, and on working holiday visas in BC, as their usual place of residence is outside of Canada. All inferences, opinions, and conclusions drawn in this manuscript are those of the authors, and do not reflect the opinions or policies of the Data Stewards.

### Study design

We report cross-sectional data describing SARS-CoV-2 testing, positivity, and COVID-19 primary care service use in BC from January 1, 2020, to July 31, 2021, stratified by immigration status (Citizen, Permanent Resident, Temporary Resident). We plot the rate of people tested and the rate of people confirmed positive for COVID-19 by week from April 19, 2020, to July 31, 2021, across immigration groups.

### Measures

Immigration status was collected from MSP registration data. People registering for MSP are required to provide documentation confirming eligible status in Canada (25). People with *citizenship* include both people born in Canada or to Canadian parents and people who immigrated and subsequently provided documentation confirming citizenship. People with *permanent residency* include economic and family class immigrants, as well as resettled refugees and successful asylum (30) or Humanitarian and Compassionate applicants. People with *temporary status* include people with work permits (including Temporary Foreign Workers), study permits, refugee claimants and convention refugees whose refugee claim was accepted but who do not yet have permanent residency. Where people held multiple statuses, we assigned them to the status held longest.

The MSP registration form contains a variable labeled “Gender” with the options “M” and “F” provided. Whether responses reflect gender, sex assigned at birth or legal sex cannot be determined. Age was calculated as of January 1, 2021. Neighbourhood income quintile was determined based on census enumeration area of residence, assigned using the Postal Code Conversion File (PCCF+) (31). We used the Statistics Canada Statistical Area Classification Metropolitan Influences Zones to group metropolitan areas (census metropolitan areas), small urban areas (census agglomerations) and rural/remote settings (areas with strong to no metropolitan influence) (32). We also report regional health authority of residence and the number of days enrolled between January 2020 and March 2021.

We determined the proportion of people who received one or more SARS-CoV-2 tests and the proportion of people with one or more positive tests in BC within the study period from the SARS-CoV-2 testing file. Access to any COVID-19 primary care is defined as one or more outpatient visit (location as office, long-term care, home, virtual) with a family doctor where the ICD9 diagnosis code submitted was C19 and/or where fee items specific to COVID-19 office visits (T13701 with test, T13702 without test) were billed to MSP. COVID-19 primary care visits could be for suspected or confirmed COVID-19 and so we report the percentage for all people, not only those with a positive SARS-CoV-2 test. Finally, we plot the number of SARS-CoV-2 tests and the number of positive tests per 100,000 people per week over the study period.

Where people had more than one test or positive tests within the week we counted only one. The denominator for all analysis includes people enrolled at any time during the study period. We have data on the number of days enrolled in MSP within the study period, but not the dates coverage started or stopped, so we cannot determine who was enrolled within each week.

### Analysis

We report outcomes as numbers and percentages by immigration group, and stratified by sex/gender, age, income quintile, rurality, and health authority. We plotted the weekly number of individuals tested and the number of individuals confirmed positive for SARS-CoV-2 per 100,000 population from April 19, 2020 to July 31, 2021 across immigration groups.

## Results

Between January 2020 and March 2021, 4,146,593 people with citizenship, 914,089 people with permanent residency and 212,215 people with temporary status registered for MSP (Table 1). Among people with temporary status, 52.1% were marked as “M”, compared to 46.4% of people with permanent residency and 50.1% of people with citizenship. Higher percentages of people with temporary status were in the 20–39-year-old age category (74.4% compared with 32.8% of people with permanent residency and 24.4% of people with citizenship), reflecting people with study and work permits. Higher percentages of people with temporary status lived in the lowest income neighbourhoods (26.5%, followed by 23.7% with permanent residency and 18.5% with citizenship). The percentage of people living in metropolitan centres was higher among people with both temporary status (87.6%) and permanent residency (90.0%) compared to people with citizenship (62.5%). Almost half of people with temporary status (47.3%) and permanent residency (50.4%) lived in the Fraser Health Authority, compared with 33.7% of people with citizenship. Mean days registered for MSP within the 456-day study period was lower among people with temporary status (355.8 days) compared with people with permanent residency (447.8 days) or citizenship (447.3 days).

**Table 1.** Characteristics of people registered for MSP who hold citizenship, permanent residency, and temporary status in British Columbia, January 2020 to July 2021, N (%)

|  | **Citizenship** | **Permanent residency** | **Temporary status** |
| --- | --- | --- | --- |
| **Total number of people** | 4,146,593 (78.6) | 914,089 (17.3) | 212,215 (4.0) |
| **Assigned sex/ legal sex/ gender\*** |  |  |  |
| F | 2,070,832 (49.9) | 490,227 (53.6) | 101,579 (47.9) |
| M | 2,075,661 (50.1) | 423,861 (46.4) | 110,636 (52.1) |
| **Age group** |  |  |  |
| 0-19 | 888,225 (21.4) | 69,124 (7.6) | 30,420 (14.3) |
| 20-39 | 1,010,046 (24.4) | 299,794 (32.8) | 157,899 (74.4) |
| 40-59 | 1,024,528 (24.7) | 337,746 (36.9) | 22,847 (10.8) |
| 60+ | 1,223,793 (29.5) | 207,425 (22.7) | 1,049 (0.5) |
| Mean age (SD) | 42.8 (24.2) | 45.6 (18.4) | 27.4 (9.4) |
| **Neighbourhood income quintile** |  |  |  |
| Lowest | 765,757 (18.5) | 216,854 (23.7) | 56,237 (26.5) |
| 2nd | 779,535 (18.8) | 205,271 (22.5) | 50,335 (23.7) |
| Middle | 829,954 (20.0) | 182,241 (19.9) | 42,859 (20.2) |
| 4th | 878,632 (21.2) | 157,811 (17.3) | 29,196 (13.8) |
| Highest | 813,501 (19.6) | 138,289 (15.1) | 25,193 (11.9) |
| Missing | 79,214 (1.9) | 13,623 (1.5) | 8,395 (4.0) |
| **Rurality** |  |  |  |
| Metropolitan | 2,591,249 (62.5) | 822,735 (90.0) | 185,817 (87.6) |
| Small urban | 934,157 (22.5) | 54,603 (6.0) | 17,157 (8.1) |
| Rural | 584,785 (14.1) | 32,577 (3.6) | 6,313 (3.0) |
| Missing | 36,402 (0.9) | 4,174 (0.5) | 2,928 (1.4) |
| **Health authority** |  |  |  |
| Interior | 790,147 (19.1) | 45,242 (4.9) | 16,048 (7.6) |
| Fraser | 1,396,969 (33.7) | 460,658 (50.4) | 100,363 (47.3) |
| Vancouver Coastal | 849,800 (20.5) | 328,578 (35.9) | 72,107 (34.0) |
| Island | 796,718 (19.2) | 62,072 (6.8) | 15,679 (7.4) |
| Northern | 281,425 (6.8) | 15,024 (1.6) | 5,439 (2.6) |
| Missing | 31,534 (0.8) | 2,515 (0.3) | 2,579 (1.2) |
| Mean days registered for MSP<br>within study period (SD)** | 447.3 (47.5) | 447.8 (44.6) | 355.8 (120.5) |

\*100 people with citizenship and <5 people with permanent residency had unknown sex/gender
\*\*The maximum is 456 (366 in 2020 and 90 in 2021, 2020 was a leap year)

The percent of people who ever received a SARS-CoV-2 test was lowest among people with temporary status (24.1%) followed by people with permanent residency (26.0%) and citizenship (28.1%) (Table 2). This pattern of lower testing among people with temporary status, followed by permanent residency, and citizenship, was observed across almost all population characteristics. Those with permanent residency had a slightly lower percentage of SARS-CoV-2 testing than those with temporary status among people over age 60. Within rural/remote settings a slightly higher percentage of people with temporary status received a SARS-Cov-2 test (21.6%) compared with people with permanent residency (20.6%) and citizenship (20.8%).

**Table 2.** COVID-19 testing, infection, and primary care use among people registered for MSP who hold citizenship, permanent residency, and temporary status in British Columbia, January 2020 to July 2021, N (%)

|  | Had one or more SARS-CoV-2 test |  |  | Had one or more SARS-CoV-2 positive result |  |  | Had one or more COVID-19 primary care visit |  |  |
| --- | --- | --- | --- | --- | --- | --- | --- | --- | --- |
|  | Citizenship | Permanent residency | Temporary status | Citizenship | Permanent residency | Temporary status | Citizenship | Permanent residency | Temporary status |
| **Total** | 1,163,694 (28.1) | 237,363 (26.0) | 51,090 (24.1) | 88,646 (2.1) | 36,918 (4.0) | 10,341 (4.9) | 182,304 (4.4) | 47,845 (5.2) | 5,526 (2.6) |
| **Assigned sex/ legal sex/ gender** |  |  |  |  |  |  |  |  |  |
| F | 624,407 (30.2) | 131,253 (26.8) | 24,970 (24.6) | 43,459 (2.1) | 18,957 (3.9) | 4,205 (4.1) | 106,415 (5.1) | 28,405 (5.8) | 2,969 (2.9) |
| M | 539,285 (26.0) | 106,110 (25.0) | 26,120 (23.6) | 45,187 (2.2) | 17,961 (4.2) | 6,136 (5.5) | 75,889 (3.7) | 19,440 (4.6) | 2,557 (2.3) |
| **Age group** |  |  |  |  |  |  |  |  |  |
| 0-19 | 245,370 (27.6) | 14,839 (21.5) | 4,127 (13.6) | 19,395 (2.2) | 2,341 (3.4) | 698 (2.3) | 29,889 (3.4) | 1,827 (2.6) | 352 (1.2) |
| 20-39 | 370,491 (36.7) | 97,504 (32.5) | 41,883 (26.5) | 30,994 (3.1) | 14,260 (4.8) | 8,517 (5.4) | 49,035 (4.9) | 16,205 (5.4) | 4,323 (2.7) |
| 40-59 | 292,280 (28.5) | 86,994 (25.8) | 4,874 (21.3) | 22,736 (2.2) | 14,211 (4.2) | 1,095 (4.8) | 52,176 (5.1) | 19,637 (5.8) | 820 (3.6) |
| 60+ | 255,553 (20.9) | 38,026 (18.3) | 206 (19.6) | 15,521 (1.3) | 6,106 (2.9) | 31 (3.0) | 51,204 (4.2) | 10,176 (4.9) | 31 (3.0) |
| **Neighbourhood income quintile** |  |  |  |  |  |  |  |  |  |
| Lowest | 210,765 (27.5) | 55,427 (25.6) | 14,098 (25.1) | 18,041 (2.4) | 9,606 (4.4) | 2,978 (5.3) | 31,746 (4.1) | 11,372 (5.2) | 1,539 (2.7) |
| 2nd | 216,203 (27.7) | 56,738 (27.6) | 13,295 (26.4) | 17,531 (2.2) | 10,332 (5.0) | 3,174 (6.3) | 34,541 (4.4) | 11,949 (5.8) | 1,417 (2.8) |
| Middle | 236,279 (28.5) | 48,504 (26.6) | 10,255 (23.9) | 18,198 (2.2) | 7,648 (4.2) | 2,110 (4.9) | 38,224 (4.6) | 10,297 (5.7) | 1,242 (2.9) |
| 4th | 258,496 (29.4) | 41,866 (26.5) | 6,919 (23.7) | 18,268 (2.1) | 5,511 (3.5) | 1,161 (4.0) | 39,358 (4.5) | 7,956 (5.0) | 704 (2.4) |
| Highest | 227,807 (28.0) | 32,145 (23.2) | 5,446 (21.6) | 15,204 (1.9) | 3,519 (2.5) | 814 (3.2) | 36,199 (4.4) | 5,811 (4.2) | 526 (2.1) |
| Missing | 14,144 (17.9) | 2,683 (19.7) | 1,077 (12.8) | 1,404 (1.8) | 302 (2.2) | 104 (1.2) | 2,236 (2.8) | 460 (3.4) | 98 (1.2) |
| **Rurality** |  |  |  |  |  |  |  |  |  |
| Metropolitan | 811,802 (31.3) | 218,053 (26.5) | 46,431 (25.0) | 68,214 (2.6) | 35,114 (4.3) | 9,545 (5.1) | 122,493 (4.7) | 43,695 (5.3) | 4,833 (2.6) |
| Small urban | 227,432 (24.3) | 12,054 (22.1) | 3,097 (18.1) | 13,109 (1.4) | 1,145 (2.1) | 555 (3.2) | 33,918 (3.6) | 2,136 (3.9) | 316 (1.8) |
| Rural | 121,657 (20.8) | 6,721 (20.6) | 1,363 (21.6) | 7,121 (1.2) | 620 (1.9) | 223 (3.5) | 25,352 (4.3) | 1,916 (5.9) | 351 (5.6) |
| Missing | 2,803 (7.7) | 535 (12.8) | 199 (6.8) | 202 (0.6) | 39 (0.9) | 18 (0.6) | 541 (1.5) | 98 (2.3) | 26 (0.9) |
| **Health authority** |  |  |  |  |  |  |  |  |  |
| Interior | 191,672 (24.3) | 10,187 (22.5) | 3,279 (20.4) | 11,310 (1.4) | 963 (2.1) | 485 (3.0) | 23,545 (3.0) | 1,376 (3.0) | 180 (1.1) |
| Fraser | 482,286 (34.5) | 140,318 (30.5) | 27,877 (27.8) | 46,582 (3.3) | 26,719 (5.8) | 7,269 (7.2) | 76,444 (5.5) | 29,642 (6.4) | 3,149 (3.1) |
| Vancouver Coastal | 257,065 (30.3) | 73,428 (22.3) | 16,651 (23.1) | 20,219 (2.4) | 8,450 (2.6) | 2,205 (3.1) | 43,046 (5.1) | 14,456 (4.4) | 1,772 (2.5) |
| Island | 176,678 (22.2) | 10,818 (17.4) | 2,318 (14.8) | 4,482 (0.6) | 384 (0.6) | 146 (0.9) | 25,629 (3.2) | 1,655 (2.7) | 222 (1.4) |
| Northern | 54,641 (19.4) | 2,451 (16.3) | 835 (15.4) | 5,970 (2.1) | 391 (2.6) | 224 (4.1) | 13,328 (4.7) | 691 (4.6) | 190 (3.5) |
| Missing | 1,352 (4.3) | 161 (6.4) | 130 (5.0) | 83 (0.3) | 11 (0.4) | 12 (0.5) | 312 (1.0) | 25 (1.0) | 13 (0.5) |

The percent of people with one or more positive SARS-Cov-2 tests was highest among people with temporary status (4.9%) followed by permanent residency (4.0%) and citizenship (2.1%). Differences in the percent of people with one or more positive tests by immigration group were greater among people marked as “M”, where 5.5% of people with temporary status had one or more positive tests, compared with 4.2% of people with permanent residency, and 2.2% of people with citizenship. The pattern of higher percentages of people who tested positive for SARS-Cov-2 among people with temporary status, followed by permanent residency, and then citizenship was observed across all income quintiles and metropolitan, urban, and rural settings. Across the entire study population, people ages 20-39, in lower income neighbourhoods, and in metropolitan centres (especially Fraser Health Authority) had higher percentages of positive tests. Within these categories, people with temporary status still had the highest percentages of positive tests.

Despite having the highest percentage of people with positive SARS-Cov-2 tests, the percentage of people with temporary status who had a COVID-19 related primary care visit was lower (2.6%) than both people with permanent residency (5.2%) and citizenship (4.4%). Differences were particularly apparent by sex/gender. While the percentage of people with positive SARS-Cov-2 tests was highest among people marked as “M” with temporary status (5.5%), access to primary care was lowest in this group (2.3%).

Examining patterns in testing over time shows that people with citizenship accessed more testing in the period preceding the second wave, and in between the second and third waves, though access to testing was similar at the peaks of wave 2 and 3 (Figure 1). Disparities in positive tests between groups were apparent in both waves 2 and 3 (Figure 2).

**Figure 1.**
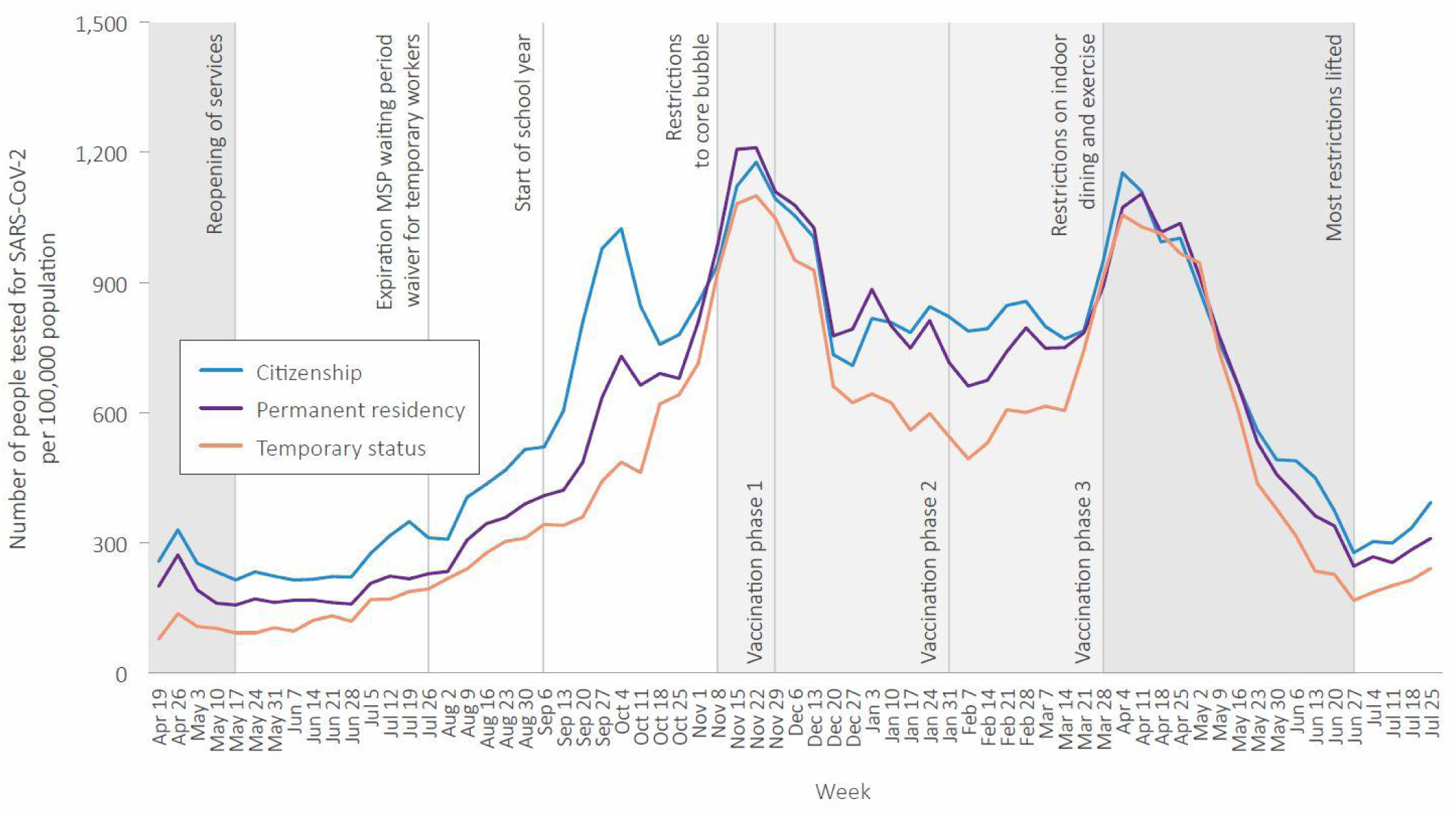
Number of people tested for SARS-CoV-2 per 100,000 population per week, by immigration status, April 2020 to July 2021.

**Figure 2.**
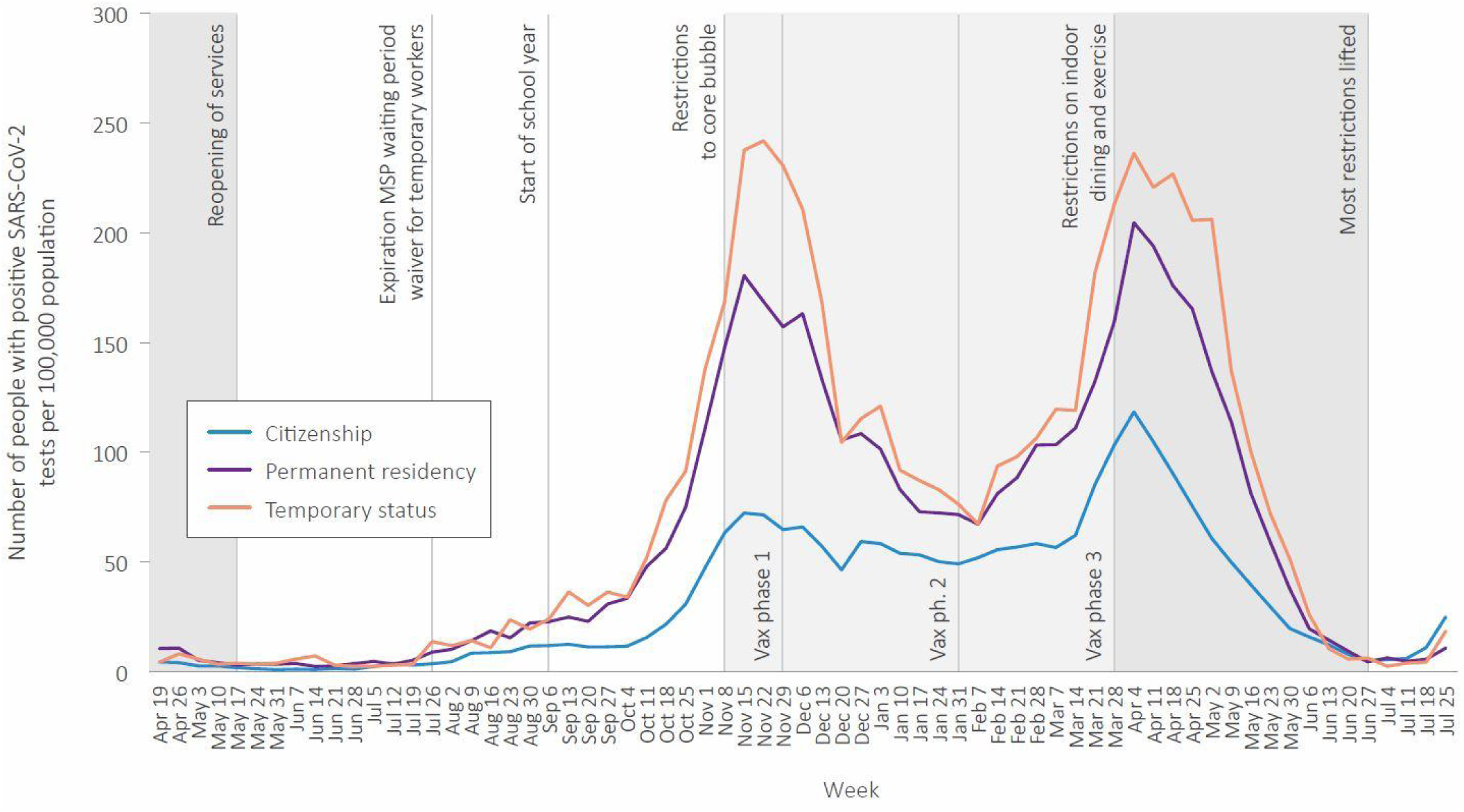
Number of people with positive SARS-CoV-2 tests per 100,000 population per week, by immigration status, April 2020 to July 2021.

## Interpretation

This study documents stark inequities in both risk of COVID-19 and access to health care, finding that temporary immigration status corresponds to higher test positivity for SARS-CoV-2, but lower access to testing and COVID-19 related primary care. This pattern is persistent by sex/gender, age group, neighborhood income quintile, health authority, and in both metropolitan and small urban settings.

More positive tests among people marked as “M” with temporary status and permanent residency, combined with lower access to primary care may suggest gendered facets of COVID-19 management, though we cannot measure gender directly due to the way the health system collects this data. Similarly, lower testing rates but higher rates of laboratory confirmed COVID-19 infection, as well as delayed testing were observed among presumed males in Ontario (33,34). In our study, higher testing and visits for COVID-19 related primary care among people with temporary status in rural/remote settings may reflect targeted testing and outreach in agricultural workplaces where people in the Temporary Foreign Worker Program are employed. However, differences in test positivity observed in Wave 2 and then Wave 3 (Fig. 2) in all geographic regions suggest targeted workplace outreach was insufficient to address dramatic differences in exposure risk and access to health services shaped by temporary immigration status.

Previous reporting has identified interconnected effects of poverty (3,35), poor housing conditions (36), occupational conditions (37), language barriers (38), systemic racism (1), and policing (39) as placing people in circumstances of higher risk of COVID-19 transmission, morbidity and mortality (40–42). People with temporary immigration status experience a convergence of some or all of these determinants, constructing a context of precarity with a profound health impact (43,44) reaching beyond health insurance coverage. For people whose temporary immigration status ends and renders them uninsured, these health impacts are magnified (45). This interaction of poverty and immigration status appears in our results, where there was increasing burden of COVID-19 with decreasing neighborhood incomes for all groups; yet, people with temporary immigration status were both more likely to live in lower income neighborhoods and experience the highest SARS-CoV-2 test positivity in those neighborhood income quintiles. The converging effects of poverty, racism, and control of movement are consistent with the history of settler colonialism and racial capitalism in the Canadian state formation (23). Canada’s immigration policy for temporary residents constrains their movement and health while benefiting from their labour and economic contributions (46). This historic and ongoing asymmetry of benefit underlies the expansion of temporary immigration programs over the past four decades, and is persistently challenged by temporary and precarious migrants advocating to uphold their human and labour rights (46). Indigenous peoples and Black communities with histories of enslavement have been here long before the border currently enforced under federal immigration policy, and continue to struggle against a contested citizenship and inequitable health outcomes (47–50).

This analysis has limitations. People who are excluded from access to provincial health insurance are excluded from this analysis. Risk of SARS-CoV-2 exposure and barriers to testing and health care may be even higher for people without health insurance in Canada, similar to uninsured people in the United States (51). People with temporary immigration status are more likely to be racialized within Canada, a dimension of COVID-19 risk observed elsewhere that cannot be measured in these data (2,52). We are similarly unable to directly measure living arrangements or employment, and income is a neighbourhood-level measure. People with temporary immigration status were not typically registered for the entire study period. This may partially explain the gap in access to COVID-19 testing and primary care for this group, though rates of positive SARS-COVo-2 tests would be even higher if the denominator was limited to people currently registered. We are missing people who registered for MSP late in the study period, as registry data was only available up to March 31, 2021. Immigration status is collected only at time of registration or renewal so may not reflect current status. The citizenship and permanent residency categories are heterogeneous, and both include people with a history of immigration. Each group contains important differences that determine circumstances of increased COVID-19 risk, for example, many disabled people experienced a higher burden of COVID-19, and may be more likely to be categorized as people with citizenship in these data as people with disabilities are excluded from most immigration pathways (53,54). Within the category of permanent residency there are economic and family class immigrants as well as refugees. Circumstances of COVID-19 risk and access to services may differ substantially within this group (2). Our analysis is not intended to examine differences between immigrants and non-immigrants defined by country of birth, but to focus on immigration status currently held. The ways in which status shapes access to services are determined by policy and within the capacity of governments to change.

People with temporary immigration status in BC experience higher SARS-CoV-2 test positivity and lower access to testing and primary care. Interwoven immigration, health and occupational policies place people in circumstances of higher health risk and will continue to amplify harms of the COVID pandemic unless all levels of government take responsibility. Extending permanent residency status to all immigrants residing in Canada and decoupling access to health care from immigration status are policies that could reduce precarity due to temporary immigration status.

## Supporting information

Strobe Checklist

COI disclosure

## Data Availability

The data that support the findings of this study are approved for use by data stewards and accessed through a process managed by Population Data BC. The data sets used for this study will be archived, and requests for access to them in the context of verification of study findings can be made to PopData (https://www.popdata.bc.ca/data_access). We are not permitted to share the research extract used in this analysis with other researchers.

## Box 1.

Citizenship, permanent and temporary status in Canada

**Canadian citizenship\*** - is granted to people born within the federal borders, who go through the formal naturalization process, or to people born to or adopted by at least one parent with Canadian citizenship. People with this status have the right to enter, remain in and leave Canada, access to the full range of rights outlined in the Canadian Charter of Rights and Freedoms, and exclusively the right to vote (55).

**Permanent residency** - is granted to people who have applied and been accepted through a variety of programs including: economic class, family class, and business class immigrants, as well as resettled refugees (government assisted, privately sponsored, and blended visa office-referred), protected persons granted permanent resident status on the basis of a well-founded fear of returning to their country of origin, and successful asylum or Humanitarian and Compassionate applicants. People with permanent residency must maintain physical residency terms, and have access to most social benefits of Canadian citizens, including health care coverage, and ability to travel and work throughout Canada. They have the option of applying for Canadian citizenship after meeting length of stay, knowledge, and official language requirements (24,56).

**Temporary status\*\*** - describes people from other countries (“foreign nationals”) who have been authorized to be in Canada for a temporary period, and includes international students, temporary workers, visitors, refugee claimants (i.e., asylum seekers) and sponsored family members. Access to health and social services is variable and dependent on provincial, municipal, local, and individual contexts. For example, some students and workers may be eligible for provincial health insurance, refugee claimants are eligible for federal health insurance, and visitors may have private insurance or pay out of pocket for health services (57,58). Local community-organized supports, along with Community Health Centres and other clinics in various places provide health care to people regardless of immigration status.

*We note this is separate from citizenship or membership in Indigenous nations, who have their own determining processes based on customary law and tradition; and, that Indigenous nations have a specific legal relationship with the federal government of Canada.

**This group falls into the larger category of precarious immigration status in Canada, which includes undocumented people who are often those in the above categories who continue to live and work in Canada beyond the expiration of their temporary documentation, and whose access to health and social services is limited.

**Appendix 1.** Characteristics of people who hold temporary status in British Columbia, January 2020-March 2021, N(%) except where indicated

|  | Student visa | Work permit | Refugee claimant | Convention refugee |
| --- | --- | --- | --- | --- |
| **Total number of people** | 108,479 | 97,789 | 2,550 | 3,397 |
| **Assigned sex/**<br>**legal sex/ gender** |  |  |  |  |
| M | 55,215 (50.9) | 43,522 (44.5) | 1,204 (47.2) | 1,638 (48.2) |
| F | 53,264 (49.1) | 54,267 (55.5) | 1,346 (52.8) | 1,759 (51.8) |
| **Age** |  |  |  |  |
| 0-19 | 28,545 (26.3) | 183 (0.2) | 816 (32.0) | 876 (25.8) |
| 20-39 | 77,333 (71.3) | 78,207 (80.0) | 948 (37.2) | 1,411 (41.5) |
| 40-59 | 2,552 (2.4) | 18,678 (19.1) | 712 (27.9) | 905 (26.6) |
| 60+ | 49 (0.0) | 721 (0.7) | 74 (2.9) | 205 (6.0) |
| **Neighbourhood**<br>**income quintile** |  |  |  |  |
| Lowest | 27,826 (25.7) | 26,614 (27.2) | 723 (28.4) | 1,074 (31.6) |
| 2nd | 25,761 (23.7) | 23,079 (23.6) | 623 (24.4) | 872 (25.7) |
| Middle | 22,419 (20.7) | 19,323 (19.8) | 538 (21.1) | 579 (17.0) |
| 4th | 13,923 (12.8) | 14,389 (14.7) | 379 (14.9) | 505 (14.9) |
| Highest | 13,420 (12.4) | 11,206 (11.5) | 248 (9.7) | 319 (9.4) |
| Missing | 5,130 (4.7) | 3,178 (3.2) | 39 (1.5) | 48 (1.4) |
| **Rurality** |  |  |  |  |
| Metropolitan | 95,265 (87.8) | 84,734 (86.6) | 2,500 (98.0) | 3,318 (97.7) |
| Small urban | 9,854 (9.1) | 7,219 (7.4) | 27 (1.1) | 57 (1.7) |
| Rural | 1,756 (1.6) | 4,539 (4.6) | 10 (0.4) | 8 (0.2) |
| Missing | 1,604 (1.5) | 1,297 (1.3) | 13 (0.5) | 14 (0.4) |
| **Health authority** |  |  |  |  |
| Interior | 9,024 (8.3) | 6,933 (7.1) | 50 (2.0) | 41 (1.2) |
| Fraser | 49,268 (45.4) | 47,230 (48.3) | 1,682 (66.0) | 2,183 (64.3) |
| Vancouver Coastal | 36,689 (33.8) | 33,623 (34.4) | 759 (29.8) | 1,036 (30.5) |
| Island | 9,212 (8.5) | 6,302 (6.4) | 44 (1.7) | 121 (3.6) |
| Northern | 2,793 (2.6) | 2,633 (2.7) | 7 (0.3) | 6 (0.2) |
| Missing | 1,493 (1.4) | 1,068 (1.1) | 8 (0.3) | 10 (0.3) |

**Appendix 2.** Number of testing episodes (and percent positivity) by immigration group

|  | **Citizenship** | **Permanent residency** | **Temporary status** |
| --- | --- | --- | --- |
| **Total Testing Episodes** | **1,808,952** | **364,819** | **69,213** |
| **Total Positive** | **93,405 (5.2)** | **39,481 (10.8)** | **10,926 (15.8)** |
| **Assigned sex/ legal sex/ gender** |  |  |  |
| F | 45,520 (4.6) | 20,152 (9.6) | 4,436 (12.9) |
| **M** | **47,885 (5.9)** | **19,329 (12.4)** | **6,490 (18.6)** |
| **Age group** |  |  |  |
| 0-19 | 19,792 (5.7) | 2,426 (12.4) | 735 (13.9) |
| 20-39 | 32,094 (5.4) | 15,100 (9.9) | 8,993 (15.8) |
| 40-59 | 23,914 (5.3) | 15,074 (11.2) | 1,165 (17.8) |
| **60+** | **17,605 (4.3)** | **6,881 (11.7)** | **33 (11.2)** |
| **Income quintile** |  |  |  |
| Lowest | 19,217 (5.7) | 10,298 (12.0) | 3,158 (16.5) |
| 2nd | 18,544 (5.5) | 11,032 (12.5) | 3,346 (18.6) |
| Middle | 19,192 (5.2) | 8,185 (10.9) | 2,229 (16.0) |
| 4th | 19,118 (4.8) | 5,906 (9.2) | 1,226 (13.1) |
| Highest | 15,857 (4.6) | 3,735 (7.8) | 860 (11.6) |
| **Missing** | **1,477 (6.7)** | **325 (8.2)** | **107 (7.6)** |
| **Rurality** |  |  |  |
| Metropolitan | 72,239 (5.5) | 37,604 (11.1) | 10,099 (16.0) |
| Small urban | 13,609 (4.1) | 1,202 (6.9) | 579 (14.7) |
| Rural | 7,346 (4.4) | 634 (6.9) | 228 (13.1) |
| **Missing** | **211 (4.7)** | **41 (5.5)** | **20 (7.8)** |
| **Health authority** |  |  |  |
| Interior | 11,630 (4.2) | 998 (6.9) | 500 (11.9) |
| Fraser | 49,923 (6.2) | 28,853 (12.9) | 7,762 (20.3) |
| Vancouver Coastal | 21,048 (5.1) | 8,817 (8.2) | 2,261 (9.9) |
| Island | 4,635 (1.9) | 402 (2.7) | 157 (5.4) |
| Northern | 6,081 (8.5) | 400 (12.7) | 232 (23.4) |

## Acknowledgements

We wish to acknowledge community partners who have provided advice on the project and to thank Dawn Mooney for preparation of Figures 1 and 2.

